# Systems Biology and Machine Learning Decode an Immunometabolic Signature for Post-Thrombotic Syndrome

**DOI:** 10.64898/2026.02.09.26345941

**Authors:** Ke Chen, Xuefeng Tian, Yang Ding, Zijian Dong, Ran Tao, Yifan Fan, Zhiyong Chen, Binshan Zha, Xiaoqiang Li, Wendong Li

## Abstract

**Objective:** Post-thrombotic syndrome (PTS), a common complication of deep vein thrombosis, lacks objective diagnostic biomarkers and its molecular mechanisms remain poorly understood. This study aimed to identify plasma biomarkers and clarify pathways using integrated multi-omics and machine learning.

**Methods:** Proteomic and metabolomic profiling of 75 PTS patients and 75 controls was performed. Differential expression analysis, pathway enrichment, and protein-metabolite network analysis were conducted. A multi-algorithm machine learning with 8 feature selection methods prioritized biomarkers. Validations and 14 models were assessed.

**Results:** 1,104 proteins and 1,891 metabolites were differentially expressed. Citrate cycle and unsaturated fatty acid biosynthesis were enriched. Three proteins, namely DIP2B, KNG1, and SUCLG2, were consistently selected as core biomarkers. All of these proteins were significantly downregulated in PTS and externally validated. A random forest model utilizing these proteins achieved an accuracy of 97.7% in independent testing, with SUCLG2 being the most influential predictor.

**Conclusion:** This study identifies a novel three - protein biomarker panel for the diagnosis of PTS and reveals an immunometabolic axis in the pathogenesis of PTS, which links inflammatory regulation with mitochondrial energy metabolism. These findings provide valuable insights into the development of diagnostic tools and targeted therapeutic approaches.

## Introduction

Deep vein thrombosis (DVT) is a prevalent vascular disorder globally, with post-thrombotic syndrome (PTS) being one of its most common and severe long-term complications^1–3^. As a chronic, disabling clinical syndrome, PTS is characterized by pain, heaviness, edema, skin pigmentation, and even venous ulcers in the affected limb. It not only severely impairs patients’ quality of life but also imposes a substantial socioeconomic burden^4^. Currently, the diagnosis of PTS primarily relies on clinical scoring systems (e.g., the Villalta score) and lacks objective biological indicators. This subjectivity leads to diagnostic heterogeneity and delays, precluding effective early intervention. More critically, there remains a dearth of reliable clinical tools to predict the progression of DVT to PTS. Thus, identifying objective and precise biomarkers for PTS diagnosis and early risk prediction has emerged as a pressing scientific priority in this field, which is pivotal for enabling individualized PTS management and improving patient outcomes^5, 6^.

In recent years, while progress has been made in elucidating the pathogenesis of PTS, it is generally acknowledged that PTS is closely associated with multiple factors, including venous wall injury, persistent thrombus residue, inflammatory responses, venous valve insufficiency, and tissue fibrosis^7, 8^. However, the specific regulatory mechanisms of the underlying molecular networks remain poorly understood. Regarding biomarker exploration, previous studies have reported potential associations between PTS and inflammatory or coagulation-related indices such as D-dimer, C-reactive protein (CRP), and interleukins^9–12^. Nevertheless, these single biomarkers are limited by low specificity and sensitivity, hindering their precise application in complex clinical settings. With the rapid advancement of high-throughput detection technologies, proteomics and metabolomics have provided robust tools for systematically deciphering the molecular landscape of diseases. Proteomics directly reflects the terminal functional states of genes, whereas metabolomics, positioned downstream of biological information flow, most directly mirrors the phenotypic and pathophysiological status of organisms. Their integration can bridge the gap from genome to phenotype, enabling a panoramic revelation of the intrinsic mechanisms governing PTS onset and progression from the dual perspectives of “functional execution” and “terminal effect.” However, research integrating proteomics and metabolomics for PTS biomarker discovery remains absent, restricting our comprehensive understanding of the disease from a systems biology standpoint.

To address these limitations, this study innovatively employed an integrated research strategy. We collected peripheral blood samples from 75 patients with definitively diagnosed PTS and 75 healthy controls, followed by parallel high-throughput proteomic and metabolomic profiling. This design allowed us to simultaneously acquire large-scale protein and metabolite expression data for the first time in PTS research, establishing a solid data foundation for systems biology analyses. Confronted with the resulting high-dimensional and complex multi-omics datasets, traditional statistical methods are often insufficient. Therefore, we incorporated multiple advanced machine learning algorithms (e.g., LASSO regression, random forest, support vector machine) and leveraged their robust feature selection and pattern recognition capabilities to efficiently and accurately screen core biomarker panels with optimal diagnostic and predictive value from the vast molecular information. This approach not only promises to construct a diagnostic model with performance far superior to that of single markers but also facilitates in-depth elucidation of the potential pathogenesis of PTS by analyzing the biological pathways enriched by these key proteins and metabolites—such as inflammatory signal transduction, immune response, energy metabolism, and oxidative stress—thereby providing a theoretical basis for the subsequent identification of novel therapeutic targets.

## Methods

### Patient selection

A total of 75 patients with PTS from the Affiliated Drum Tower Hospital of Nanjing University Medical School were included in the experimental group. The diagnosis was based on scoring, CTV or lower extremity venography, with scores all above 10 points, indicating moderate to severe PTS. They were diagnosed 1-2 years after DVT treatment. Meanwhile, 75 patients without PTS after DVT treatment during the same period were included in the control group. Subsequently, peripheral blood from a total of 150 patients was collected. After centrifugation, the plasma was extracted for proteomics and metabolomics studies. Two prospective validation cohorts were from the Jiangbei Campus of the Affiliated Drum Tower Hospital of Nanjing University Medical School (57 cases of PTS and 57 controls) and the First Affiliated Hospital of Anhui Medical University (88 cases of PTS and 88 controls). This study was approved by the Ethics Committee of the Affiliated Drum Tower Hospital of Nanjing University Medical School (Ethics number: 2024-294-02, 2024-05-22) and the First Affiliated Hospital of Anhui Medical University(Ethics number: PJ-2024-11-23, 2024-11-23). All participants gave written informed consent.

Inclusion criteria for the PTS group: 1. Confirmed lower extremity deep vein thrombosis and treated within 6 months to 2 years, with diagnosis based on lower extremity deep vein angiography; 2. Score greater than 10 points, and imaging examination (CTV or angiography) indicated PTS. Exclusion criteria for the PTS group: 1. Recurrence of lower extremity deep vein thrombosis; 2. No anticoagulant treatment after confirmed DVT; 3. Severe liver or kidney dysfunction or long-term use of hormones; 4. Active stage of malignant tumors or active stage of rheumatic diseases. The control group included patients who were diagnosed with lower extremity deep vein thrombosis and treated within 6 months to 2 years, with a score less than 5 points.The baseline data of the included patients are shown in **Table 1**.

**Table 1.** Baseline of patients enrolled in PTS and non-PTS group.

| name | levels | Non-PTS (N=75) | PTS (N=75) | P value |
| --- | --- | --- | --- | --- |
| Length_of_hospital_stay | Median (IQR) | 7.0 (5.0 to 11.0) | 9.0 (6.0 to 12.5) | 0.108 |
| Age | Median (IQR) | 64.0 (50.5 to 74.5) | 67.0 (56.5 to 73.5) | 0.307 |
| Sex | Men | 42 (56%) | 38 (50.7%) | 0.513 |
|  | Woman | 33 (44%) | 37 (49.3%) |  |
| BMI | Mean ± SD | 23.4 ± 3.5 | 24.3 ± 3.6 | 0.138 |
| Smoking | No | 73 (97.3%) | 59 (78.7%) | <0.001 |
|  | Yes | 2 (2.7%) | 16 (21.3%) |  |
| Drinking | No | 68 (90.7%) | 67 (89.3%) | 0.785 |
|  | Yes | 7 (9.3%) | 8 (10.7%) |  |
| Location_of_the_thrombus | Iliac vein | 4 (5.3%) | 8 (10.7%) | 0.168 |
|  | Femoral vein | 46 (61.3%) | 51 (68%) |  |
|  | Popliteal vein, intermuscular vein | 25 (33.3%) | 16 (21.3%) |  |
| History_of_surgery | No | 55 (73.3%) | 51 (68%) | 0.473 |
|  | Yes | 20 (26.7%) | 24 (32%) |  |
| History_of_tumors | No | 67 (89.3%) | 51 (68%) | 0.001 |
|  | Yes | 8 (10.7%) | 24 (32%) |  |
| Radiotherapy | No | 71 (94.7%) | 66 (88%) | 0.147 |
|  | Yes | 4 (5.3%) | 9 (12%) |  |
| chemotherapy | No | 71 (94.7%) | 68 (90.7%) | 0.347 |
|  | Yes | 4 (5.3%) | 7 (9.3%) |  |
| fracture | No | 66 (88%) | 61 (81.3%) | 0.257 |
|  | Yes | 9 (12%) | 14 (18.7%) |  |
| Hypertension | No | 58 (77.3%) | 52 (69.3%) | 0.268 |
|  | Yes | 17 (22.7%) | 23 (30.7%) |  |
| coronary_heart_disease | No | 72 (96%) | 72 (96%) | 1.000 |
|  | Yes | 3 (4%) | 3 (4%) |  |
| DM | No | 62 (82.7%) | 59 (78.7%) | 0.535 |
|  | Yes | 13 (17.3%) | 16 (21.3%) |  |
| Homocysteinemia | No | 75 (100%) | 75 (100%) | 1.000 |
|  | Yes | 0 (0%) | 0 (0%) |  |
| CKD | No | 71 (94.7%) | 61 (81.3%) | **0.012** |
|  | Yes | 4 (5.3%) | 14 (18.7%) |  |
| Thrombophilia | No | 65 (86.7%) | 65 (86.7%) | 1.000 |
|  | Yes | 10 (13.3%) | 10 (13.3%) |  |
| pregnant | No | 73 (97.3%) | 75 (100%) | 0.155 |
|  | Yes | 2 (2.7%) | 0 (0%) |  |
| DVT_Family_history | No | 75 (100%) | 75 (100%) | 1.000 |
|  | Yes | 0 (0%) | 0 (0%) |  |
| Immune_diseases | No | 74 (98.7%) | 74 (98.7%) | 1.000 |
|  | Yes | 1 (1.3%) | 1 (1.3%) |  |
| WBC | Median (IQR) | 7.3 (5.5 to 9.3) | 6.4 (5.4 to 8.9) | 0.675 |
| Neutrophil | Median (IQR) | 4.7 (3.5 to 7.1) | 4.3 (3.3 to 6.7) | 0.625 |
| Lymphocyte | Median (IQR) | 1.4 (1.2 to 1.9) | 1.4 (1.1 to 1.9) | 0.462 |
| Macrophage | Median (IQR) | 0.5 (0.3 to 0.6) | 0.5 (0.3 to 0.7) | 0.373 |
| PT | Median (IQR) | 13.2 (12.3 to 14.1) | 13.8 (12.7 to 15.1) | **0.010** |
| APTT | Median (IQR) | 34.7 (29.8 to 39.4) | 37.1 (33.2 to 42.5) | **0.019** |
| INR | Median (IQR) | 1.0 (1.0 to 1.1) | 1.1 (1.0 to 1.2) | **0.016** |
| Hb | Mean ± SD | 114.4 ± 22.2 | 118.4 ± 17.7 | 0.223 |
| PLT | Median (IQR) | 201.0 (164.0 to 260.5) | 210.0 (170.0 to 251.0) | 0.621 |
| CR | Median (IQR) | 63.0 (53.9 to 77.0) | 61.0 (51.5 to 79.0) | 0.919 |
| BUN | Median (IQR) | 5.3 (4.1 to 6.9) | 4.9 (3.9 to 6.5) | 0.729 |
| ALT | Median (IQR) | 22.0 (15.0 to 31.5) | 28.0 (18.5 to 33.5) | 0.068 |
| AST | Median (IQR) | 20.0 (15.0 to 27.0) | 20.0 (18.0 to 27.0) | 0.262 |
| TB | Median (IQR) | 11.3 (8.6 to 15.7) | 13.4 (10.4 to 17.2) | 0.052 |
| GFR | Median (IQR) | 101.0 (92.0 to 116.9) | 100.0 (86.0 to 111.8) | 0.444 |
| D_Dimer | Median (IQR) | 0.7 (0.6 to 6.1) | 8.4 (1.3 to 13.3) | <0.001 |

### Plasma collection and processing

Human peripheral blood was drawn into EDTA-coated tubes. Plasma was isolated through a two-step centrifugation protocol: an initial low-speed spin to remove blood cells, followed by a high-speed centrifugation to eliminate platelets and other debris. The purified plasma was immediately aliquoted and frozen at −80°C to preserve protein integrity for subsequent proteomic analysis.

### Proteomics

Plasma samples were centrifuged at 14,000g for 20 min, and the supernatant was subjected to protein quantification via BCA assay. Aliquots of 15 µg protein were combined with 5X loading buffer, denatured by boiling, and resolved on 4%–20% SDS-PAGE gels at 180 V for 45 min, followed by Coomassie Brilliant Blue R-250 staining. For sample sets larger than six, equal amounts of protein from individual samples were pooled to create a quality control Pool. All samples, including the Pool, were digested using the FASP method. The resulting peptides were desalted using C18, lyophilized, and reconstituted in 0.1% formic acid for concentration measurement at OD280. After spiking with iRT peptides, the samples were analyzed by nano-LC (Vanquish Neo) coupled with an Astral mass spectrometer in positive ion mode. Full-scan MS1 spectra were acquired from 380 – 980 m/z at a resolution of 240,000, followed by data-independent acquisition (DIA) MS2 with 299 windows and 25 eV collision energy. DIA-NN software was employed for data processing against the Uniprot database, allowing one missed cleavage, fixed carbamidomethylation of cysteine, variable modifications of methionine oxidation and protein N-terminal acetylation, and a protein false discovery rate (FDR) below 1%.

### Metabolomics

For metabolomic analysis, metabolites were extracted from 100μL of plasma using 400 μL of pre-cooled methanol:acetonitrile (3:1, v/v) solution. The mixture was vortexed, sonicated, and incubated at 4°C. After centrifugation, the supernatant was dried under vacuum and reconstituted in 50% methanol. The analysis was performed using a UHPLC system (HSS T3 column) coupled to a Thermo QE HF-X mass spectrometer. Chromatographic separation was achieved with a 12-minute gradient elution at a flow rate of 0.3 mL/min. MS data were acquired in both positive and negative ESI modes under DDA, with a scan range of 70-1050 m/z. Raw data files were processed using Compound Discoverer 3.3 for peak picking, alignment, and compound identification against databases (mzCloud, KEGG, HMDB, etc.). Subsequent data analysis was conducted using tools within the Linux environment, including R and Python.

### Multi-Omics Analysis Pipeline

#### Proteomics Data Processing and Differential Expression Analysis

Raw protein quantification values were log₂-transformed to stabilise variance and approximate a normal distribution prior to statistical testing. Differentially expressed proteins (DEPs) between patients with PTS and controls (non-PTS after DVT) were identified using independent samples *t*-tests, with statistical significance set at *p* < 0.05. Fold change (FC) was calculated as the ratio of mean protein abundance between the two groups. Proteins meeting both *p* < 0.05 and |log₂FC| > 1 were classified as DEPs.

#### Metabolomics Data Processing and Differential Expression Analysis

For metabolomics profiling, discrimination between PTS patients (n =75) and healthy controls (n=75) was first assessed using orthogonal projections to latent structures discriminant analysis (OPLS-DA). The OPLS-DA model generated variable importance in projection (VIP) scores to quantify each metabolite’s contribution to group separation. Independent samples *t*-tests were performed to obtain metabolite-level *p*-values, and fold change was calculated to determine the direction of regulation (FC > 1 for upregulation; FC < 1 for downregulation). Metabolites were defined as differentially expressed metabolites (DEMs) when they satisfied three criteria simultaneously: VIP > 1, |log₂FC| > 1, and *p* < 0.05.

#### KEGG Pathway Enrichment Analysis

DEPs were subjected to KEGG pathway enrichment analysis using the clusterProfiler package (default parameters). For metabolites with valid KEGG Compound IDs, enrichment analysis was performed through the MetaboAnalyst platform. Fisher’s exact test was used to assess enrichment significance, and the Benjamini–Hochberg method was applied to control for false discovery. Enriched pathways for DEPs were defined as those with adjusted *p* < 0.05, whereas enriched pathways for DEMs were considered significant at *p* < 0.05, reflecting platform-specific output formats.

#### Protein–Metabolite Interaction Network Construction

To investigate functional relationships between DEPs and DEMs, interaction information was retrieved from the STITCH database, a comprehensive resource of predicted and experimentally validated protein–chemical associations. Protein–metabolite interaction pairs were extracted together with their combined scores, which represent the confidence of each interaction. These data were used to construct an integrated interaction network linking proteins and metabolites associated with the enriched pathways.

#### Network Module Detection and Functional Annotation

Community detection within the interaction network was performed using the cluster_edge_betweenness function in the *igraph* R package. This method, also known as the Girvan–Newman algorithm, identifies network modules by iteratively removing edges with the highest betweenness centrality. For each detected module, we enumerated the KEGG pathways associated with the proteins contained in that module. Pathways with the high recurrence were considered the dominant functional categories of the corresponding module, thereby enabling interpretation of major biological themes connecting proteomic and metabolomic alterations in PTS.

### Machine Learning Analysis

#### Initial Feature Screening Using Multiple Algorithms

To identify key protein features associated with PTS, we implemented a multi-algorithm screening strategy using the caret, catboost, and lightgbm packages in R. Eight supervised learning algorithms—Random Forest (RF), Gradient Boosting Machine (GBM), Lasso regression, Partial Least Squares (PLS), Linear Discriminant Analysis (LDA), LightGBM (LGBM), CatBoost (CatB), and Decision Tree (DT)—were applied. Each model was trained using a targeted set of 75 proteins, comprising 17 DEPs associated with Biosynthesis of unsaturated fatty acids and the Citrate cycle (TCA cycle) and 18 additional “connection” DEPs identified through protein–metabolite network analysis. These 35 proteins constituted the expression matrix used to estimate variable importance.

#### Feature Importance Evaluation and Consensus Selection

Model interpretability was performed using the explain() function from the DALEX package, which calculates feature importance for each trained model. Proteins with zero or negligible importance scores were removed. Proteins identified as important in at least seven of the eight models were retained as consensus features for the next stage of analysis. Visualisation of the feature-selection results was completed using ComplexHeatmap, UpSetR, tidyverse, and ggpubr.

#### Model Development and Hyperparameter Optimisation

For model development, all 150 samples were randomly partitioned into a training set (70%) and a test set (30%), maintaining 1:1 case–control proportions. Using the caret, catboost, and lightgbm packages, we fitted 14 machine learning classifiers with the selected protein features as predictors: Adaptive Boosting, Bayesian Generalised Linear Model, Gradient Boosting, CatBoost, Linear Discriminant Analysis, LightGBM, Lasso, Logistic Regression, k-Nearest Neighbours, Neural Network, Random Forest, Support Vector Machine with radial kernel, XGBoost, and a general Boosting method. Hyperparameters for each model were optimised using **10-fold cross-validation** on the training set, and final model performance was evaluated on the independent test set.

#### Model Performance Evaluation

Model performance metrics included accuracy, F1-score, sensitivity, specificity, positive predictive value (PPV), negative predictive value (NPV), and Youden’s index, calculated separately for the training and test sets. Models that demonstrated high discriminative ability with minimal overfitting were selected for further evaluation using calibration curves.

#### SHAP-Based Model Interpretation

To interpret the contribution of individual proteins to model predictions, we applied the kernelshap package to compute SHAP (SHapley Additive exPlanations) values. Visualisation and ranking of SHAP values were performed using the shapviz package, enabling ranking and interpretation of protein influence.

## Results

### 1. Differential Proteomic and Metabolomic Profiles in PTS

**Figure 1** illustrates the overall experimental approach. A total of 1,104 differentially expressed proteins (DEPs) were identified between PTS patients and healthy controls, of which 181 were upregulated and 923 were downregulated based on the criteria *p* < 0.05 and |log₂FC| > 1 (**Figure 2A**). KEGG pathway enrichment analysis showed that the upregulated proteins were mainly associated with immune and inflammatory processes, including Complement and coagulation cascades, Protein processing in the endoplasmic reticulum, Wnt signalling pathway, Cytokine–cytokine receptor interaction, and Neutrophil extracellular trap formation (**Figure 2B**).

**Figure 1.**
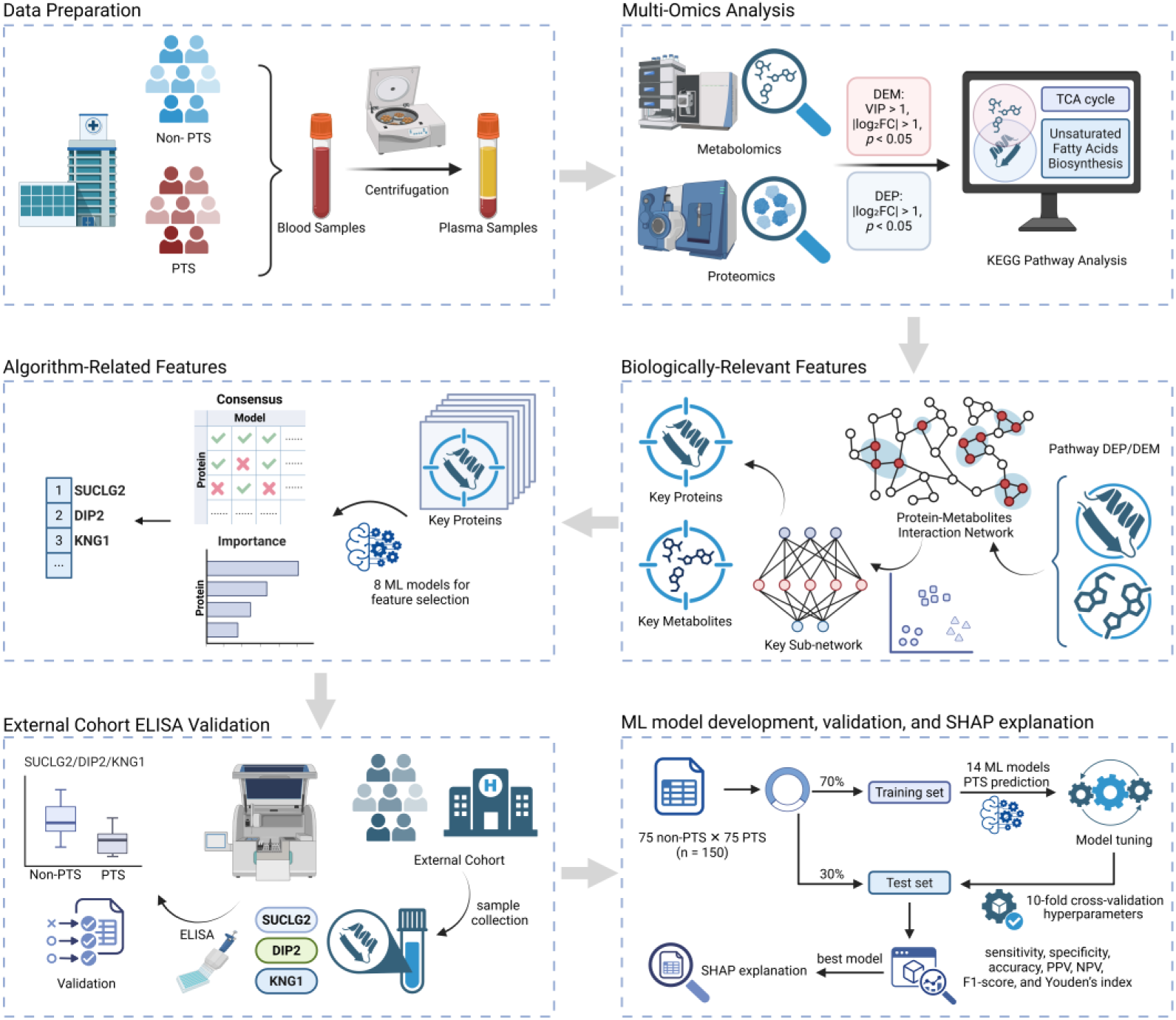
Flowchart of experimental design.

**Figure 2.**
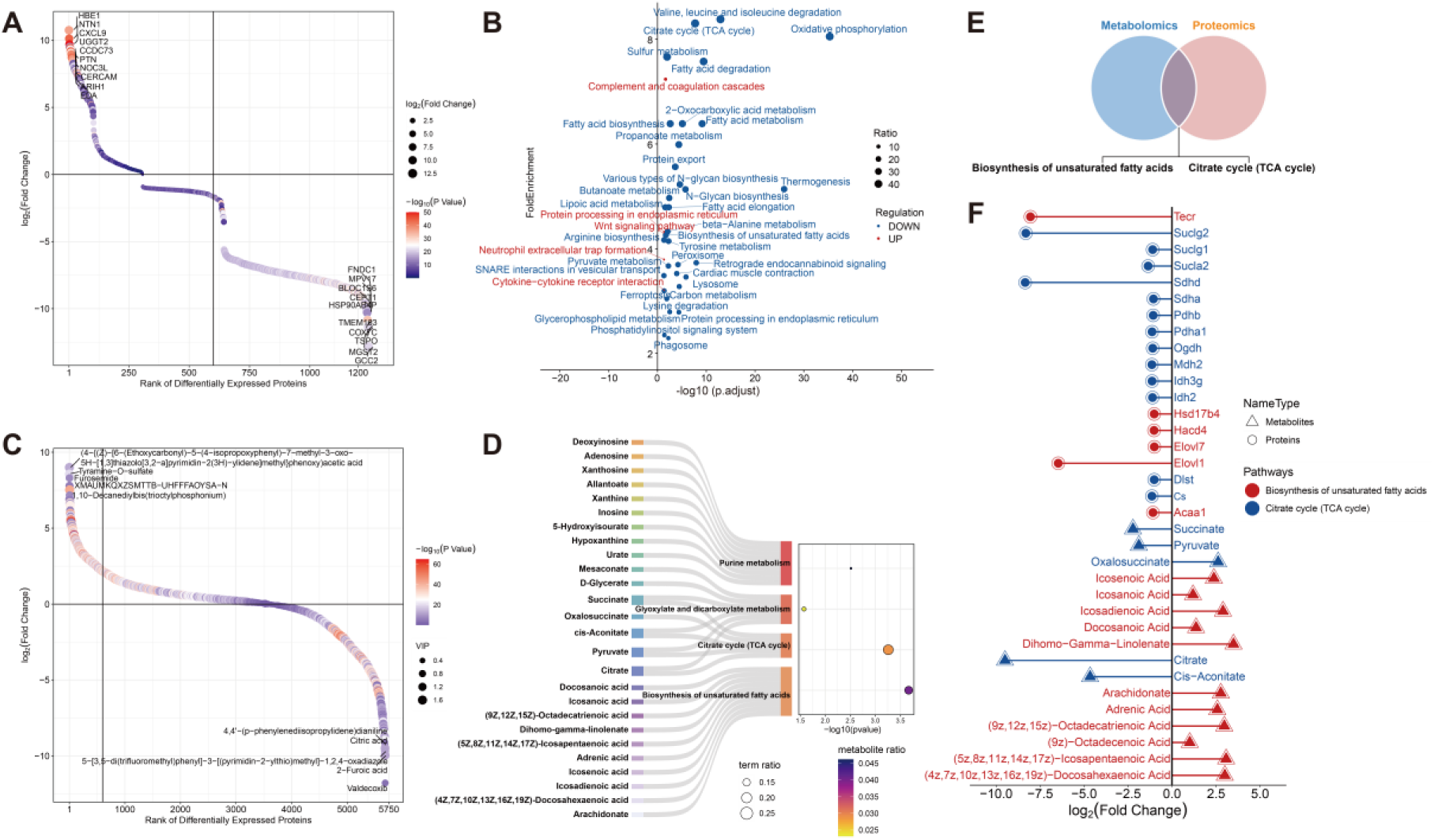
Differential proteomic and metabolomic profiles in post-thrombotic syndrome (PTS). (A) Overview of differentially expressed proteins (DEPs) identified between PTS patients (n = 75) and non-PTS (n = 75) based on the criteria *p* < 0.05 and |log₂FC| > 1. (B) KEGG pathway enrichment analysis of upregulated and downregulated DEPs. Enrichment significance for proteins was defined as adjusted *p* < 0.05. Upregulated proteins were mainly enriched in immune and inflammatory pathways, whereas downregulated proteins were predominantly enriched in mitochondrial and metabolic pathways. (C) Overview of differentially expressed metabolites (DEMs) identified between PTS patients and healthy controls using the criteria *p* < 0.05, |log₂FC| > 1, and VIP > 1. (D) KEGG pathway enrichment analysis of DEMs (significance threshold: *p* < 0.05), highlighting major metabolic pathways associated with energy production and lipid metabolism. (E) Venn diagram showing the two cross-omics overlapping KEGG pathways between proteomics and metabolomics datasets: Biosynthesis of unsaturated fatty acids and Citrate cycle (TCA cycle). (F) Key DEPs and DEMs mapped to the two shared pathways. Tecr and Dihomo-γ-linolenate showed the largest absolute fold changes within the unsaturated fatty acid biosynthesis pathway, while Suclg2 and citrate showed the most prominent decreases within the TCA cycle.

In contrast, the downregulated proteins were enriched predominantly in pathways linked to mitochondrial activity and substrate metabolism, such as Oxidative phosphorylation, Thermogenesis, Valine, leucine and isoleucine degradation, Fatty acid degradation, and Fatty acid metabolism. These pathways collectively reflect reduced energy production and impaired metabolic capacity in PTS.

Metabolomics analysis identified 1,891 differentially expressed metabolites (DEMs), including 1,230 upregulated and 661 downregulated species (*p* < 0.05, |log₂FC| > 1, VIP > 1) (**Figure 2C**). KEGG enrichment revealed four significantly enriched metabolic pathways: Purine metabolism, Glyoxylate and dicarboxylate metabolism, Citrate cycle (TCA cycle), and Biosynthesis of unsaturated fatty acids (**Figure 2D**). These pathways are central to cellular energy production, redox balance, and lipid biosynthesis, indicating broad metabolic disturbances in PTS.

An intersection analysis of protein- and metabolite-level KEGG enrichments identified two shared pathways: Biosynthesis of unsaturated fatty acids and the Citrate cycle (TCA cycle) (**Figure 2E**). Within the unsaturated fatty acid biosynthesis pathway, the protein with the largest absolute fold change was Tecr, which was markedly downregulated, while the metabolite with the highest change was Dihomo-γ-linolenate, which was strongly upregulated. For the TCA cycle, Suclg2 represented the most downregulated protein, and citrate was the most substantially decreased metabolite (**Figure 2F**). These results suggest that perturbations in energy metabolism and lipid biosynthesis may contribute to the metabolic dysfunction underlying PTS pathogenesis.

### 2. Integrated proteomic–metabolomic interaction landscape

The integrated protein–metabolite interaction network revealed twelve major functional modules, which were largely centred on energy production, material metabolism, and signal regulation. Key examples included oxidative phosphorylation, arachidonic acid metabolism, fatty acid metabolism, and the tricarboxylic acid (TCA) cycle, highlighting that these pathways form the metabolic core of post-thrombotic syndrome (PTS).

Metabolites and proteins enriched in shared KEGG pathways (**Figure 2E**) were not only clustered within the TCA cycle and unsaturated fatty acid metabolism but also extended into additional modules such as oxidative phosphorylation, arachidonic acid metabolism, general fatty acid metabolism, and cytokine–cytokine receptor interaction (**Figure 3A**). These clusters suggest that the biological disturbances in PTS are coordinated across several connected metabolic and signalling processes rather than confined to a single pathway.

**Figure 3.**
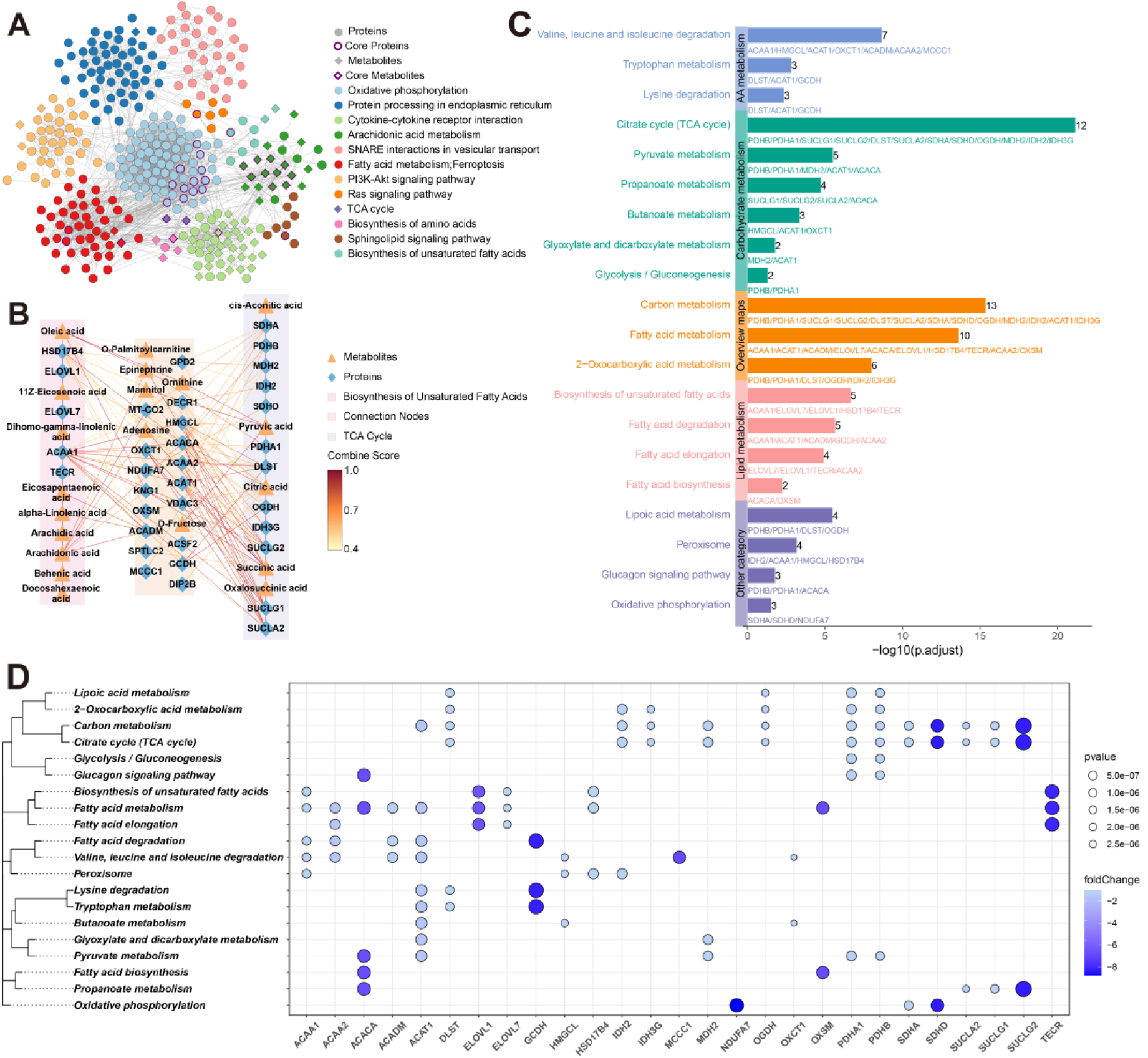
Integrated proteomic–metabolomic interaction landscape in PTS. (A) Integrated protein–metabolite interaction network constructed using the STITCH database. Proteins and metabolites involved in the two cross-omics shared pathways—Biosynthesis of unsaturated fatty acids and Citrate cycle (TCA cycle)—are highlighted. The network reveals multiple functional modules mainly associated with energy metabolism, lipid metabolism, and signal regulation. (B) Sub-network showing proteins and metabolites with direct cross-omic interactions within the two shared pathways and exhibiting high interaction strength (combined score > 0.4). (C) KEGG pathway enrichment analysis of the 35 proteins obtained by integrating interaction node proteins with pathway-enriched proteins (significance threshold: adjusted *p* < 0.05). The most significantly enriched pathways were the Citrate cycle (TCA cycle), Carbon metabolism, and Fatty acid metabolism. (D) Differentially expressed proteins (DEPs) involved in the significantly enriched pathways. Suclg2 shows the largest absolute fold change and participates in multiple energy-related pathways. ACAT1, PDHA1, PDHB, DLST, and ACAA1 are the most frequently enriched proteins.

To further define the central molecular features, we isolated proteins and metabolites that showed direct cross-omic interactions within both shared pathways and exhibited high interaction strength (combined score > 0.4). These were considered key nodes that link the two pathways at a functional level (**Figure 3B**). When we merged the interaction nodes with the pathway-enriched proteins, we obtained a set of 35 proteins, among which 27 proteins appeared in the KEGG enrichment analysis. The three most significantly enriched pathways were Citrate cycle (TCA cycle), Carbon metabolism, and Fatty acid metabolism, again reflecting a strong emphasis on energy and substrate metabolism (**Figure 3C**).

Within this set, the most strongly downregulated protein, Suclg2, had the highest absolute log fold change. Suclg2 participates in the TCA cycle, propanoate metabolism, and carbon metabolism, underscoring its central role in mitochondrial energy production. In addition, the proteins most frequently represented across enrichment analyses—ACAT1, PDHA1, PDHB, DLST, and ACAA1—serve as key metabolic enzymes positioned at crucial entry points or branch points of mitochondrial fuel processing (**Figure 3D**).

Together, these results provide an initial systems-level view of the proteomic and metabolomic alterations associated with PTS. The consistent involvement of pathways linked to unsaturated fatty acid metabolism and core energy metabolism suggests that PTS may involve a broader metabolic disruption, particularly within mitochondrial processes that govern energy generation and substrate utilisation.

### 3. Machine-learning–based feature screening identifies three key PTS-associated proteins

Feature selection was conducted on the 35-protein panel generated from pathway-enriched proteins and high-interaction node proteins. Using eight machine learning algorithms, we evaluated variable importance to identify proteins most consistently associated with PTS. Three proteins—DIP2B, SUCLG2, and KNG1—were repeatedly highlighted across the models, each being selected by at least seven algorithms (**Figure 4A**). Among them, DIP2B and SUCLG2 were identified by all eight methods, while KNG1 appeared in seven models, with the exception of the decision tree classifier.

**Figure 4.**
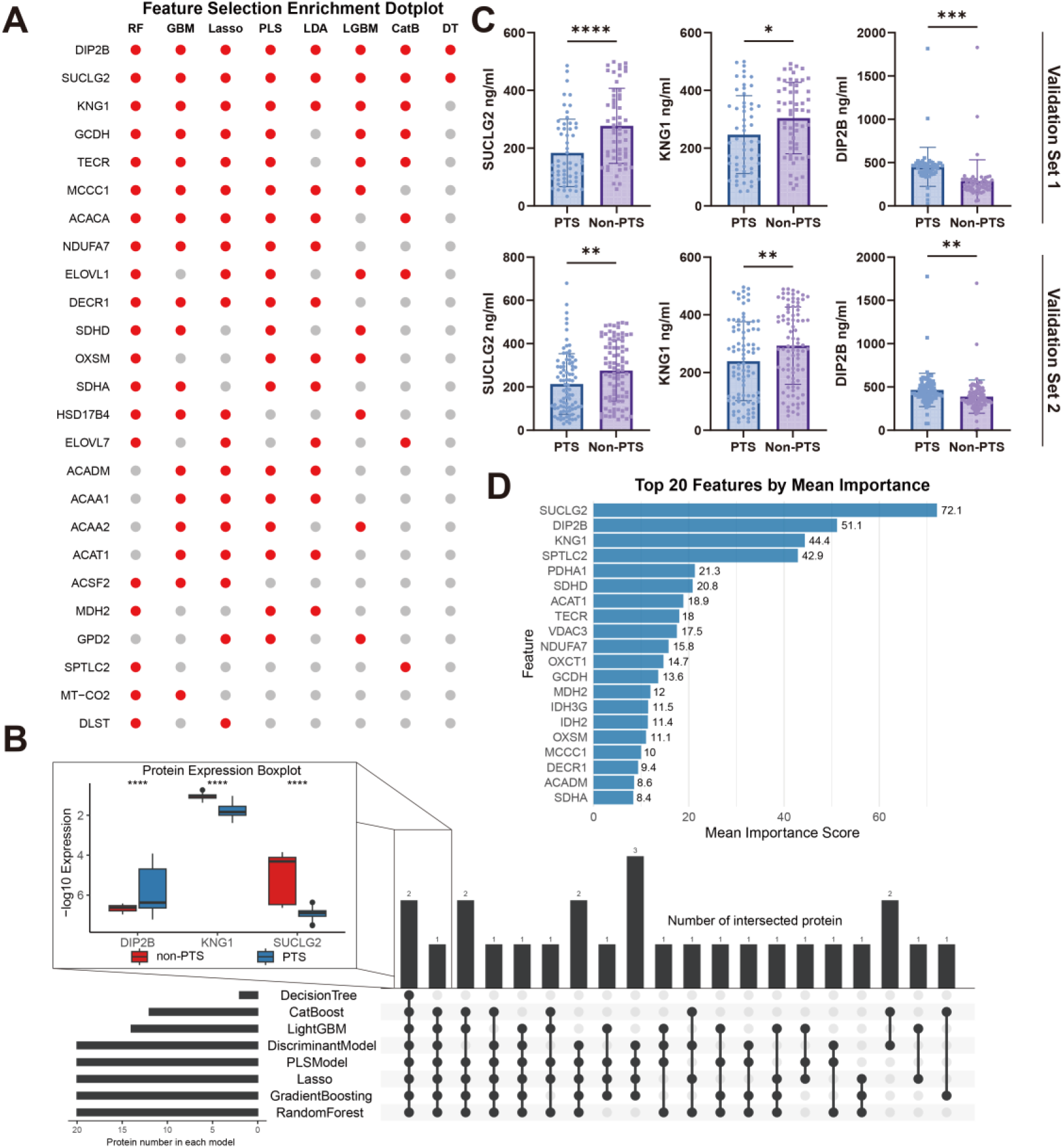
Machine-learning–based feature selection identifies three key PTS-associated proteins. (A) Consensus dot plot showing feature selection results for the 35 candidate proteins evaluated across eight machine learning algorithms. Each dot indicates that a protein was selected as an important feature by the corresponding algorithm. (B) Three proteins—SUCLG2, KNG1, and DIP2B—were consistently identified by at least seven algorithms. Their expression levels in PTS patients and non-PTS are shown as box plots. Statistical significance was assessed using independent *t*-tests (*p* < 0.05: *, < 0.01: **, < 0.001: ***, < 0.0001: ****). (C) Validation of DIP2B, SUCLG2, and KNG1 expression levels by ELISA in two independent cohorts (validation set 1: n = 57 per group; validation set 2: n = 88 per group). All three proteins showed consistent downregulation in PTS (*t*-test significance as indicated). (D) Top 20 protein features ranked by mean importance across the eight machine learning algorithms. SUCLG2 ranks highest, followed by DIP2B and KNG1.

All three proteins were significantly downregulated in patients with PTS (**Figure 4B**). To validate the proteomic observations, we measured their circulating levels using ELISA in two independent cohorts, both of which showed consistent reductions in DIP2B, SUCLG2, and KNG1, supporting the robustness of the findings (**Figure 4C**).

When ranking proteins by their average importance across the eight algorithms, SUCLG2 emerged as the top-ranked feature, followed by DIP2B and KNG1 (**Figure 4D**). Taken together, these results indicate that DIP2B, SUCLG2, and KNG1 represent the most reliable and biologically meaningful features for distinguishing individuals with PTS, and therefore were selected as the core predictors for subsequent model development.

### 4. Development and evaluation of machine-learning models for PTS prediction

To construct predictive models for PTS, the three key proteins—DIP2B, SUCLG2, and KNG1—were incorporated into 14 machine-learning algorithms (Adaptive Boosting, Bayes Method, Boosting Method, CatBoost, Discriminant Model, Gradient Boosting, Lasso, LightGBM, Logistic Model, Nearest Neighbour Method, Neural Network, Random Forest, SVM Kernel, and XGBoost). Model training was performed on the training cohort, and hyperparameters for each algorithm were tuned using 10-fold cross-validation within the *caret* framework to ensure optimal performance.

Across the training set, all models achieved high sensitivity, specificity, accuracy, PPV, NPV, F1-score, and Youden’s index, suggesting strong discriminative ability (**Figure 5A**). Performance on the independent test set remained similarly robust, with consistently high values across the same evaluation metrics (**Figure 5B**), indicating good model generalisability.

**Figure 5.**
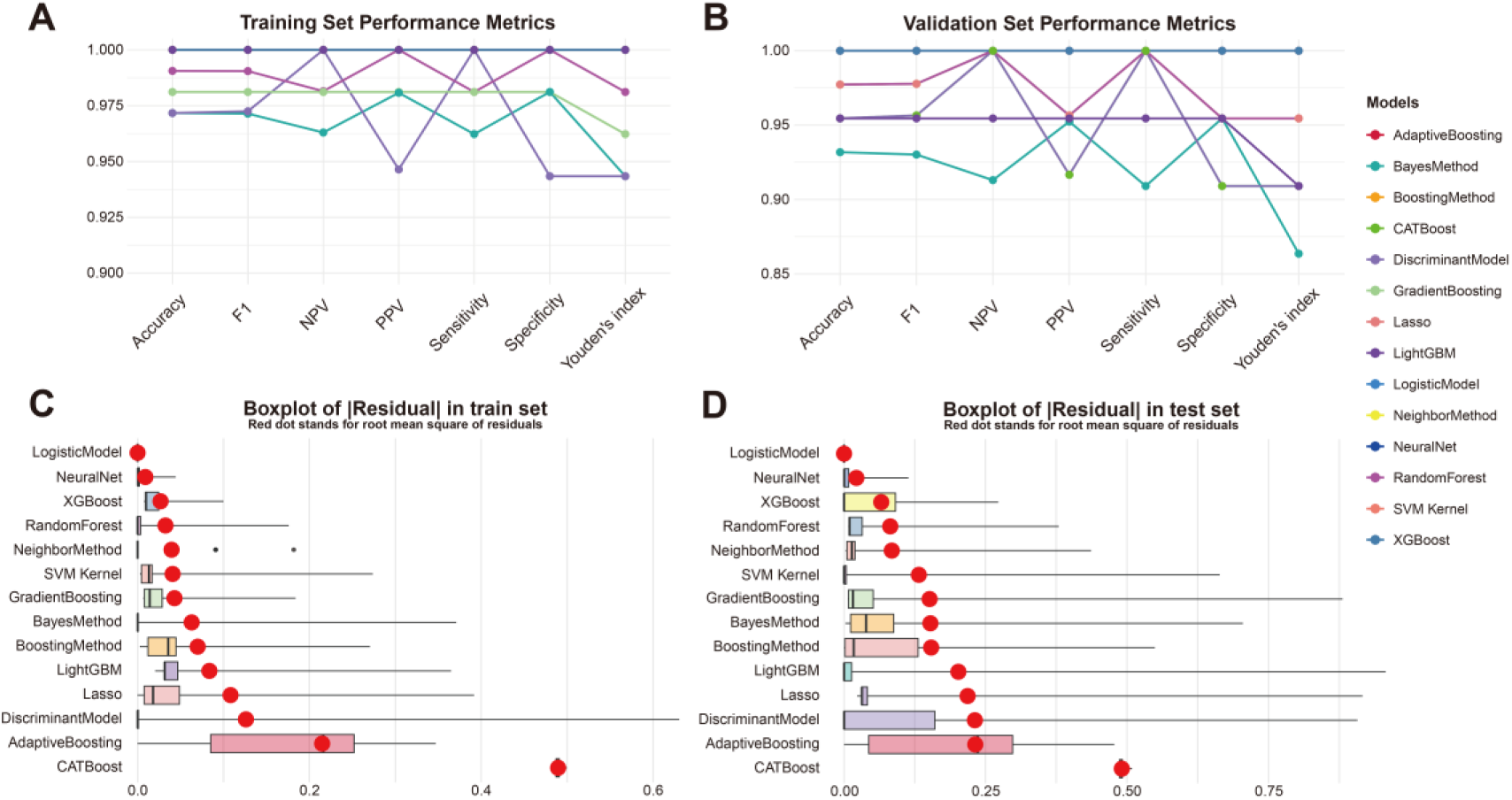
Development and evaluation of machine-learning models for PTS prediction. (A) Predictive performance of 14 machine-learning algorithms in the training cohort, evaluated using accuracy, F1-score, negative predictive value (NPV), positive predictive value (PPV), sensitivity, specificity, and Youden’s index. (B) Predictive performance of the same 14 machine-learning algorithms in the independent test cohort, showing consistently robust classification ability across all evaluation metrics. (C) Boxplot of the absolute residuals (|Residual|) for each model in the training set, illustrating model fitting performance. The five models with the lowest residuals were Logistic Model, Nearest Neighbour Method, Random Forest, XGBoost, and Neural Network. (D) Boxplot of the absolute residuals (|Residual|) for each model in the test set, demonstrating that the same five models also exhibited the most stable generalisation performance across independent data.

To compare overall performance, we examined the mean residuals of each model. The five algorithms with the lowest residuals on the training set were: Logistic Model, Nearest Neighbour Method, Random Forest, XGBoost, and Neural Network (**Figure 5C**). Importantly, the same five models also showed the smallest residuals on the test set (**Figure 5D**), highlighting their stable behaviour across datasets.

Although several models performed strongly, we noted that extremely high accuracy on the training set may signal an overfitting tendency in some models. After balancing predictive performance with model stability and interpretability, the Random Forest model was selected for subsequent interpretative analyses due to its strong cross-validated performance and lower susceptibility to overfitting relative to more complex algorithms.

### 5. Model interpretation using SHAP

Following model selection, the random forest classifier was further evaluated and interpreted using SHAP. The model demonstrated excellent discriminative performance. In the training set, all 106 samples were correctly classified, resulting in an accuracy of 100% (**Figure 6A**). In the test set, only one healthy control was incorrectly classified as PTS, giving an overall accuracy of 97.7% (**Figure 6B**). Calibration curves for both the training and test cohorts showed good alignment between predicted and observed probabilities, indicating that the model was well-calibrated and maintained stable predictive behaviour (**Figure 6C–D**).

**Figure 6.**
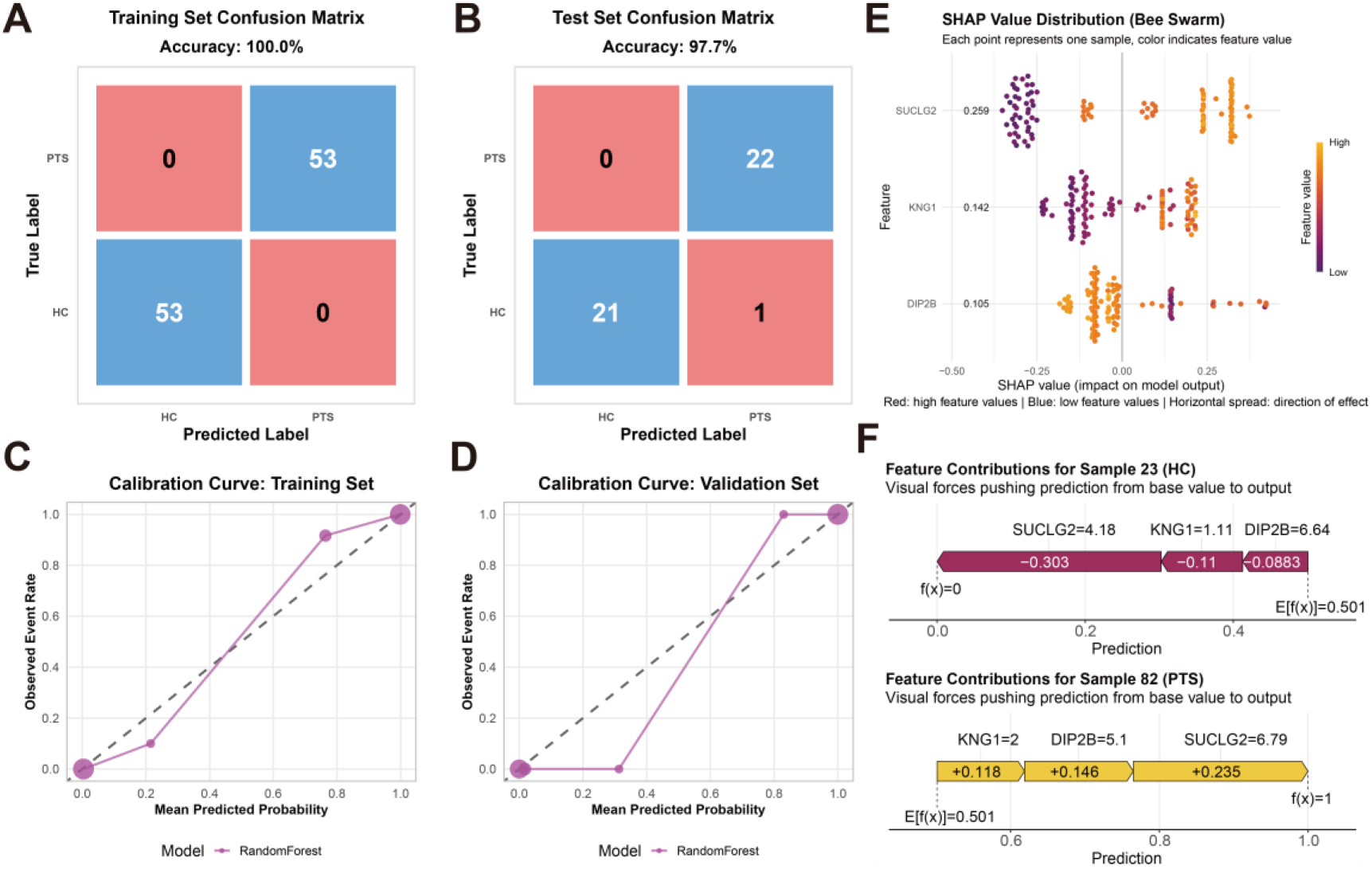
Performance evaluation and model interpretation of the random forest classifier using SHAP. (A) Confusion matrix of the training cohort demonstrating perfect classification of all 106 samples, with no misclassified cases. (B) Confusion matrix of the test cohort, showing that only one healthy control was misclassified as PTS, yielding an overall accuracy of 97.7%. (C) Calibration curve of the training cohort, illustrating excellent agreement between predicted probabilities and observed outcomes across the full probability range. (D) Calibration curve of the test cohort, confirming that the model maintained stable and reliable probability estimates in the independent validation dataset. (E) SHAP summary (bee swarm) plot showing the global importance and distribution of SHAP values for the three predictors. Each dot represents one sample, with colour indicating feature expression level. SUCLG2 exhibits the highest mean absolute SHAP value, indicating the strongest influence on model predictions, followed by KNG1 and DIP2B. (F) SHAP force plots for representative samples, illustrating how individual protein features contribute to the final prediction for a healthy control and a PTS patient. In both cases, SUCLG2 exerts the dominant directional effect on the model output, while KNG1 and DIP2B provide additional contributory effects.

To explore how individual proteins contributed to model predictions, we applied SHAP to quantify variable importance. Among the three predictors, SUCLG2 displayed the highest mean absolute SHAP value, confirming its dominant influence on classification, followed by KNG1 and DIP2B (**Figure 6E**). This ranking was consistent with the feature-importance results from the broader machine-learning screening.

To illustrate how these proteins shaped the prediction process at the individual level, we examined control and PTS using SHAP force plots. In both examples, SUCLG2 exerted the strongest directional effect on the model output, pushing predictions either towards or away from PTS depending on its expression pattern. KNG1 and DIP2B contributed to a lesser but still meaningful extent.

## Discussion

By integrating proteomic and metabolomic analyses of peripheral blood with diverse machine learning algorithms, this study successfully identified three core protein biomarkers (DIP2B, KNG1, SUCLG2) and key metabolic pathways closely associated with the development of PTS.

These findings not only provide a highly valuable biomarker panel for the objective diagnosis of PTS but also reveal, at the molecular level, that its pathophysiology may be deeply rooted in dysregulated immune-inflammatory responses and cellular energy metabolic reprogramming.

The three key proteins we identified play central roles in the inflammatory and metabolic disturbances of PTS. DIP2B, potentially expressed in immune cells such as monocytes/macrophages, may be involved in regulating inflammatory signaling pathways (e.g., NF-κB)^13, 14^. Following deep vein thrombosis, the organization and recanalization of the thrombus are accompanied by intense inflammation. Dysfunctional DIP2B could lead to uncontrolled or prolonged inflammatory responses, exacerbating damage to the venous wall and valves. Furthermore, DIP2B dysregulation might promote fibroblast activation, resulting in excessive deposition of extracellular matrix components like collagen, which reduces venous wall elasticity, causes luminal narrowing, and aggravates venous return impairment. In parallel, the observed downregulation of KNG1 underscores the significance of microvascular dysfunction in PTS^15–17^. KNG1 is the precursor of bradykinin, a core effector of the kallikrein-kinin system with potent vasodilatory, algesic, and vascular permeability-enhancing effects. Reduced KNG1 levels may reflect a failure of compensatory regulation against vascular leakage and tissue edema in the context of chronic inflammation, aligning closely with the characteristic clinical manifestations of limb swelling and pain in PTS patients^18, 19^.

To address the challenges of high dimensionality and noise inherent in high-throughput omics data, we employed 14 distinct machine learning algorithms for cross-validation and feature selection. This strategy’s primary strength lies in its ability to transcend the limitations of conventional univariate statistical analyses, enabling the robust identification of the most predictive and stable biomarker combinations from complex, non-linear variable interactions. Different algorithms—leveraging their respective principles such as L1 regularization (LASSO), Gini impurity (Random Forest), and maximum margin separation (SVM)—collectively converged on DIP2B, KNG1, and SUCLG2. This consensus significantly enhances the reliability of our findings, indicating that these biomarkers are not spuriously associated but are robustly and profoundly linked to the pathological state of PTS, thereby laying a solid foundation for developing a generalizable clinical diagnostic model.

Notably, SUCLG2 shifts the focus from classical inflammation to the emerging frontier of cellular energy metabolism^20^. SUCLG2 is a component of succinyl-CoA ligase, a key enzyme in the tricarboxylic acid (TCA) cycle that catalyzes the conversion of succinyl-CoA to succinate coupled with GTP generation^21^. Dysregulation of SUCLG2 directly impairs TCA cycle efficiency and overall mitochondrial energy production^22^. Its identification as a plasma biomarker is particularly noteworthy, suggesting a systemic reflection of altered mitochondrial energy metabolism in PTS. The chronic inflammatory state in PTS may be sustained by fundamental metabolic reprogramming, potentially through mechanisms such as aerobic glycolysis, driving immune cells toward a pro-inflammatory phenotype. These specific metabolic disturbances provide functional context for the proteomic signals^23^. TCA cycle disruption, consistent with impaired mitochondrial oxidative phosphorylation, could promote the persistence of activated immune cells and lead to tissue damage. Additionally, observed alterations in unsaturated fatty acid metabolism are highly relevant to inflammatory processes, as these lipids serve as precursors for pro-inflammatory eicosanoid signaling molecules. This metabolic shift likely contributes to the persistent vascular inflammation and oxidative stress characteristic of PTS^24, 25^. Our model positions cellular metabolic dysfunction as a potential contributor to the chronicity of PTS, rather than merely a consequence, revealing new directions for investigating metabolic pathways as therapeutic targets.

In summary, this study proposes an integrative hypothesis for PTS pathogenesis: following DVT, persistent pathological stimuli trigger vascular homeostatic imbalance, marked by KNG1 downregulation, concurrently inducing a mitochondrial energy metabolism crisis characterized by SUCLG2 dysfunction. Metabolic disturbances lead to insufficient energy supply and elevated oxidative stress, which not only directly impair vascular function but also exacerbate inflammatory cascades and tissue injury. A limitation of this study is that all findings are based on peripheral blood samples. Future work should validate the functions of these proteins and metabolic pathways within local vascular tissues using cellular and animal models, and further explore their causal relationships. Nonetheless, the biomarker panel we discovered establishes a solid foundation for developing non-invasive diagnostic tools for PTS, and the revealed metabolism-inflammation axis mechanism provides promising directions for future therapies targeting energy metabolism and regulated cell death pathways.

## Author Contribution

Ke Chen, Yang Ding, Xuefeng Tian and Zijian Dong: original manuscript writing, data analysis, picture assembly and statistical analysis.

Ke Chen, Ran Tao,Yifan Fan: Clinical data collection, application of ethical documents.

Xiaoqiang Li, Wendong Li, Zhiyong Chen, Binshan Zha: Project design and optimization, manuscript revision and optimization, funding provision and overall quality control.

## Declaration

All listed authors have reviewed the final manuscript and consent to its submission in the current form. The research described herein has not been previously published and is not under consideration for publication elsewhere, in whole or in part. Each author has contributed sufficiently to the work to be included as a co-author. The authors declare no competing interests.

## Data Availability Statement

All data have been included in the supplementary materials. If any investigators have any other needs, they can get additional data from.

## Acknowledgement

We would like to thank all the teachers and partners of Laboratory 615, Scientific Research Laboratory Building, The Affiliated Drum Tower Hospital, Nanjing University Medical School.

## Ethical approval

All procedures performed in studies involving human participants were in accordance with the ethical standards of the institutional and/or national research committee and with the 1964 Helsinki declaration and its later amendments or comparable ethical standards. All participants gave written informed consent.

This study was approved by the Ethics Committee of the Affiliated Drum Tower Hospital of Nanjing University Medical School (Ethics number: 2024-294-02, 2024-05-22) and the First Affiliated Hospital of Anhui Medical University (Ethics number: PJ-2024-11-23, 2024-11-23).

## Supplementary figures

**Supplementary Figure 1.**
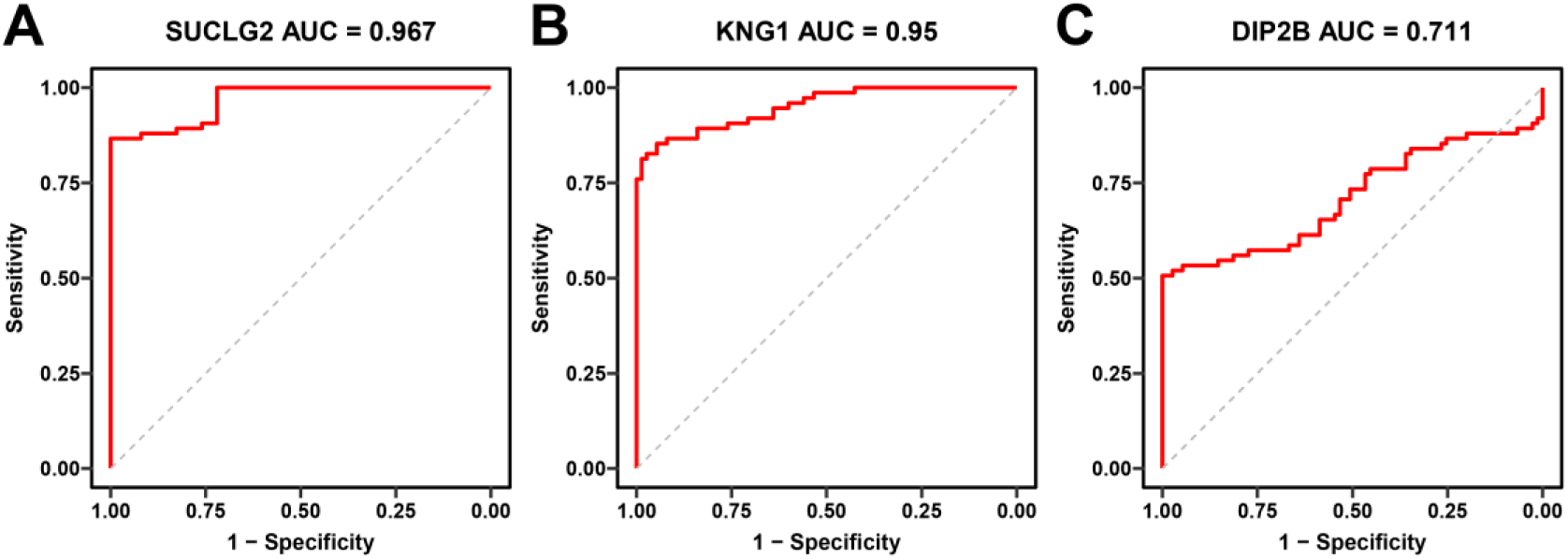
Diagnostic performance of individual protein biomarkers for PTS. Receiver operating characteristic (ROC) curves were generated using the expression levels of each individual protein to evaluate their ability to discriminate between PTS patients and healthy controls. (A) ROC curve of SUCLG2, showing excellent diagnostic performance with an area under the curve (AUC) of 0.967. (B) ROC curve of KNG1, also demonstrating strong discriminative ability with an AUC of 0.950. (C) ROC curve of DIP2B, showing moderate predictive performance with an AUC of 0.711.

**Supplementary Figure 2.**
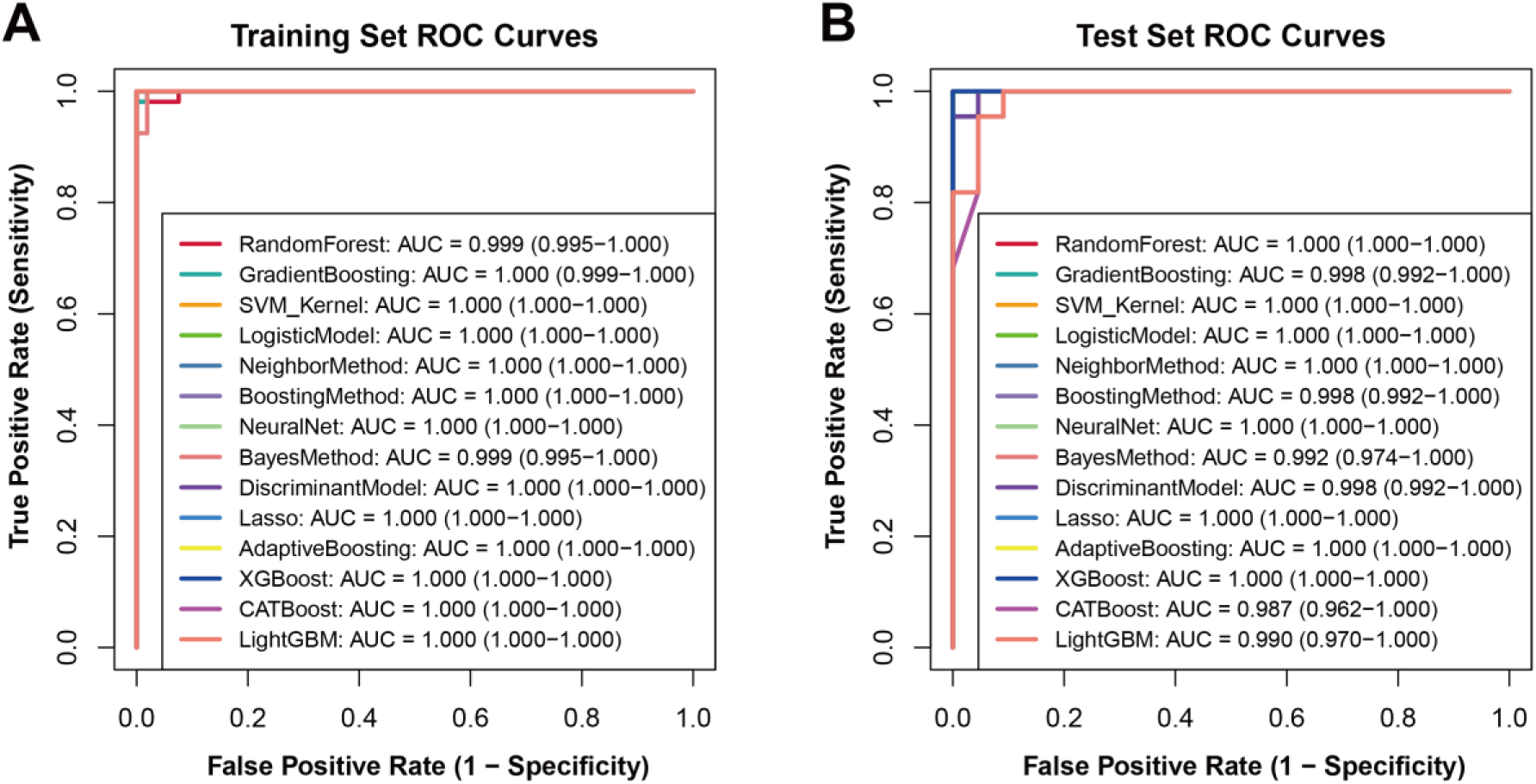
ROC curves of the 14 machine-learning models in the training and test sets. (A) ROC curves and corresponding area under the curve (AUC) values for the 14 machine-learning algorithms in the **training set**. All models exhibited near-perfect discriminative performance, with AUC values ranging from **0.999 to 1.000**, indicating excellent classification ability during model fitting. (B) ROC curves and AUC values for the same 14 algorithms in the **test set**, demonstrating consistently high predictive performance, with AUC values ranging from **0.987 to 1.000**.

